# Phantom-based quantification of the spectral accuracy in dual-layer spectral CT for pediatric imaging at 100 kVp

**DOI:** 10.1101/2022.02.27.22271573

**Authors:** Sebastian Meyer, Leening P. Liu, Harold I. Litt, Sandra S. Halliburton, Nadav Shapira, Peter B. Noël

**Affiliations:** Department of Radiology, Perelman School of Medicine, University of Pennsylvania, Philadelphia, PA 19103, USA; Philips Healthcare, Orange Village, OH 44122, USA; Department of Diagnostic and Interventional Radiology, School of Medicine & Klinikum rechts der Isar, Technical University of Munich, 81675 München, Germany

**Keywords:** Tomography, X-ray computed, Radiation dose, Phantoms, Imaging

## Abstract

**Objectives:** To determine the spectral accuracy in detector-based dual-energy CT (DECT) at 100 kVp and wide (8 cm) collimation width for dose levels and object sizes relevant to pediatric imaging.

**Methods:** A spectral CT phantom containing tissue-equivalent materials and iodine inserts of varying concentrations was scanned on the latest generation detector-based DECT system. Two 3D-printed extension rings were used to mimic varying pediatric patient sizes. Scans were performed at 100 and 120 kVp, 4 and 8 cm collimation widths, and progressively reduced radiation dose levels, down to 10% of the nominal value for the standard pediatric abdominal protocol. Virtual mono-energetic, iodine density, effective atomic number, and electron density results were quantified and compared to their expected values for all acquisition settings and phantom sizes.

**Results:** DECT scans at 100 kVp provided highly accurate spectral results; however, a size dependence was observed for iodine quantification. For the medium phantom configuration (15 cm diameter), measurement errors in iodine density, effective atomic number, and electron density (ED) were below 0.3 mg/ml, 0.2 and 1.8 %ED_water_, respectively. The average accuracy was slightly different from scans at 120 kVp; however, not statistically significant for all configurations. Collimation width had no substantial impact. Spectral results were accurate and reliable for radiation exposures down to 0.9 mGy CTDI_vol_.

**Conclusions:** Detector-based DECT at 100 kVp can provide on-demand or retrospective spectral information with high accuracy even at extremely low doses, thereby making it an attractive solution for pediatric imaging.

**Key points:**

- Dual-layer spectral CT at 100 kVp enables high-quality spectral imaging for smaller patients
- Larger collimation width does not affect the accuracy of spectral results
- Accurate and reliable spectral quantification is achievable at radiation doses down to 0.9 mGy

## Introduction

Dual-energy CT (DECT) [1–3], the first clinical realization of spectral imaging, is an imaging technology that enables unique quantification of clinically relevant materials based on their elemental composition. By utilizing data from two different x-ray spectra, DECT overcomes several limitations of conventional single-energy CT imaging. Dedicated decomposition algorithms that exploit the material-specific energy dependency of x-ray attenuation [2, 4–6] provide energy- and material-selective reconstructions. These include virtual mono-energetic images (VMIs) [7], quantitative material density assessment and virtual subtraction of clinical contrast agents, as well as direct information on electron density (ED) and effective atomic number (Z_eff_) [8].

Multiple solutions have been developed for generating spectral data based on different detection and source technologies [1, 2]. Among the commercially available DECT systems, dual-layer spectral CT has the unique property of creating the spectral separation at the detector level. This design uses a single x-ray source and two attached, but optically separated, scintillation detectors with different sensitivity profiles. While the upper (inner) layer selectively absorbs low-energy x-rays and serves as filtration, the bottom (outer) layer is more sensitive to higher energy x-rays [9, 10].

This detector-based spectral data collection provides multiple advantages. First, both spectral and conventional anatomical images are generated simultaneously for every scan. Hence, one does not need to prospectively select studies for spectral acquisitions and the original imaging workflow can be maintained. Spectral imaging has only been possible at 120 and 140 kVp, but the latest generation of detector-based DECT also enables acquisitions at 100 kVp. Detector-based spectral separation also provides perfect spatial and temporal alignment of both spectra, enabling efficient projection-space decomposition with an intrinsic reduction of beam-hardening artefacts [7] and better spectral denoising compared to image-space decomposition [9].

While the general clinical benefit of spectral information is well established [11–13], there is less evidence concerning the potential of DECT for pediatric imaging. Current applications of DECT in children include improved automated bone removal for CT angiography, assessment of lung blood volume as a surrogate for perfusion imaging, reduction of metal artifacts originating from highly-attenuating materials such as orthopedic hardware, and diagnosis of abdominal neoplasms [14–16].

One reason for the limited use of DECT in pediatric imaging are the unique requirements of this patient population. First, radiation exposure from medical procedures is a paramount concern for children and must be kept as low as reasonably achievable [17]. Due to their longer lifetime expectancy and increased radiosensitivity of their cells compared to adults, children have a higher cumulative risk of developing radiation-induced cancers. Thus, pediatric CT protocols routinely use lower x-ray beam energies of 100 kVp for abdomen and chest scans to reduce the radiation exposure while maintaining or even improving diagnostic image quality [18]. Second, young children are often unable to comply with instructions and do not remain still during extended scan times, which increases the likelihood for motion artifacts [19]. Using larger collimation widths and fast gantry rotation times, down to 0.27 seconds, allows reduction of the overall acquisition time, potentially avoiding the need for sedation. The aim of this work is a quantitative investigation of the impact of using 100 kVp tube voltage and 8 cm beam collimation width on the spectral imaging performance for pediatric imaging using a state-of-the-art dual-layer DECT system.

## Methods

### 1. Phantom configuration

In this study, we used a spectral CT phantom (QRM GmbH) with custom-made extension rings. The cylindrical 10 cm diameter phantom (Figure 1) is composed of water-equivalent plastic and contains tissue-equivalent inserts corresponding to liver, adipose, and 100 and 400 mg/ml hydroxyapatite (HA) materials. For iodine density quantification, four solid iodine inserts with properties equivalent to iodinated contrast media at concentrations of 0.5, 2, 5 and 10 mg/ml were also included. To incorporate the large variability of body size in children and adolescents and assess its impact on spectral quantification, two supplemental extension rings of 15 and 20 cm outer diameter were 3D-printed from a polylactate acid (PLA) filament. The rings were attached to the original phantom (Figure 1) and the selected diameters (10, 15 and 20 cm) reflect typical waist circumferences in children aged 2 to 18 years [20, 21].

**Figure 1.**
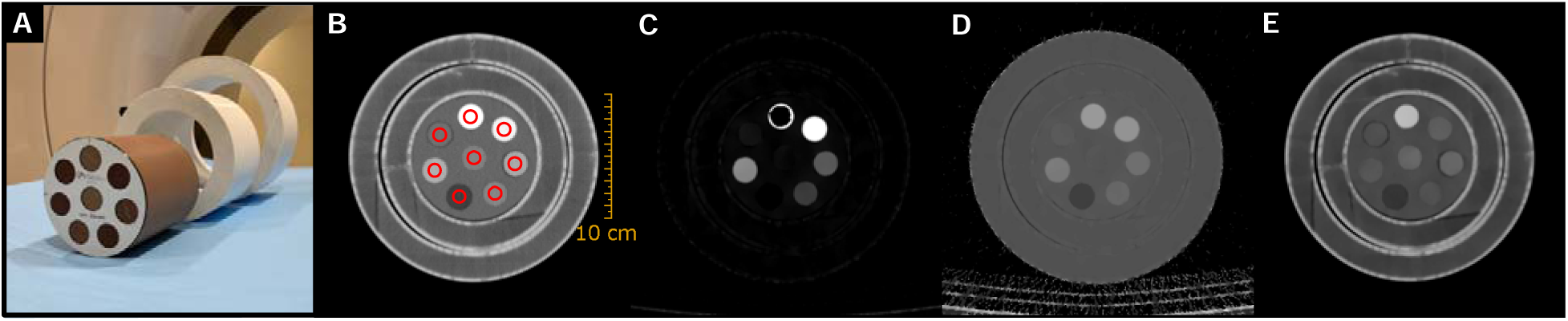
(A) Spectral CT phantom consisting of a central part with eight inserts and two removable extensions rings. (B) Virtual monoenergetic image at 70 keV (window level/width = 50/500 HU) including regions of interest for image quality evaluations, (C) iodine density map (window level/width = 5/10 mg/ml), (D) effective atomic number map (window level/width = 10/10), and (E) electron density map (window level/width = 110/50 %ED_water_) images are shown.

To evaluate the accuracy of the spectral results, the expected (i.e., ground truth) values for all inserts were calculated from the manufacturer material composition specifications. Nominal Hounsfield unit (HU) values for a virtual monoenergetic images (VMI) of energy *E* are calculated as:

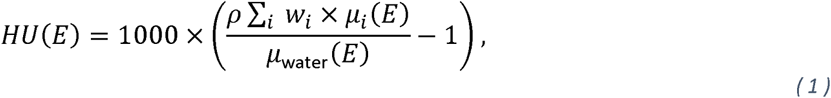

where *i* ranges over the elements composing the material and *w*_*i*_ is the normalized mass fraction of element *Z*_*i*_ with atomic mass *A*_*i*_. *ρ* represents the physical density and the mass attenuation coefficients (*μ*_*i*_ and *μ*_water_ for element *i* and water, respectively) were obtained from the NIST database [22]. The effective atomic number of an insert material is given by [8, 23]:

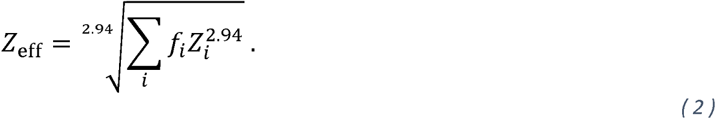

*f*_*i*_ corresponds to the fractional number of electrons of the constituent element *Z*_*i*_. The exponent 2.94 is selected based on published literature [8]. Electron density (ED) values provided by the scanner are expressed as percentage relative to water (*ED*_*w*_ *=* 3.343 × 10^23^ m^−3^) and can be calculated as:

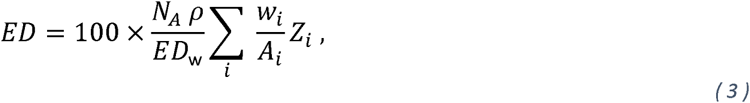

where *N*_*A*_ is Avogadro’s constant. The nominal values for all insert materials used in this work can be found in Shapira et al. [23].

### 2. Data acquisition

All acquisitions were performed on a commercial dual-layer spectral CT scanner (Spectral CT 7500, Philips Healthcare). Scans used the default pediatric (0-10 kg) abdomen protocol with 0.272 second gantry rotation time and a pitch of 1.234. Acquisitions were performed at 120 and 100 kVp tube voltages as well as 4 and 8 cm (total) collimation widths. The phantom was positioned in the iso-center and a similar positioning was maintained for all three phantom configurations. For each scenario (i.e., combination of scan parameter and phantom size), images were obtained at four different dose levels (CTDI_vol_) of 9 (default pediatric abdominal protocol), 6, 3 and 0.9 mGy. Three repetitions were performed for each scan.

The image series were reconstructed with the hybrid iterative reconstruction method iDose^4^ (spectral denoising level of 2) and a clinical standard kernel for body imaging (B). In total, 48 datasets (2 tube voltages x 4 dose levels x 2 collimation widths x 3 repetitions) were collected for each phantom configuration. We did not investigate different spectral denoising levels as they have no substantial impact on the accuracy of spectral results [8, 24]. The resulting spectral base images contain all relevant information to generate VMIs, as well as iodine density, Z_eff_, and ED images. A summary of all acquisition and reconstruction parameters is presented in Table 1.

**Table 1.** DECT acquisition and reconstruction parameters.

|  |  |
| --- | --- |
| Tube voltage [kVp] | 120, 100 |
| Collimation width [mm] | 40 (64 × 0.625), 80 (128 × 0.625) |
| Dose level (CTDI<sub>vol</sub>) [mGy] | 9, 6, 3, 0.9 |
| Tube current exposure time product [mAs] | • 120kVp/40 mm: 109, 72, 36, 11<br>• 120kVp/80 mm: 116, 78, 39, 11<br>• 100kVp/40 mm: 175, 116, 58, 17<br>• 100kVp/80 mm: 188, 125, 63, 19 |
| Pitch | 1.234 |
| Rotation time [s] | 0.272 |
| Slice thickness [mm] | 3 |
| Field-of-view [mm<sup>2</sup>] | 250×250 |
| Pixel spacing [mm] | 0.488 |

### 3. Image quality assessment

Spectral accuracy was measured using circular regions of interest (ROIs) within each cylindrical insert on 26 consecutive axial slices in the center of the phantom. For each configuration, ROIs were positioned manually on the high dose 70 keV VMIs and covered only half of the actual insert radius to avoid inaccuracies originating from partial volume effects (Figure 1). Subsequently, these ROIs were used to extract pixel data from all datasets, i.e., conventional, VMIs, iodine density, Z_eff_ and ED, and the mean value was obtained for each slice. Statistical analysis was performed using MATLAB R2021b (MathWorks). Differences in spectral quantification accuracy (i.e., mean error) for different collimation widths were compared using Mann–Whitney U tests. The Kruskal-Wallis test was used to compare different dose levels and phantom sizes. In addition, we analyzed changes in spectral quantification variation (i.e., standard deviation of the measurement error) for different dose levels using Kruskal-Wallis tests followed by a post hoc pairwise comparison with Dunn’s test (using the Šidák correction). In all cases a p-value ≤ 0.05 was considered as statistically significant.

## Results

We first present each individual spectral result at the pediatric abdominal protocol nominal radiation dose (9 mGy CTDI_vol_). The impact of dose reduction was evaluated separately in the last subsection.

### 1. Virtual mono-energetic images

The accuracy of various VMIs (from 40 keV to 200 keV) is summarized in Table 2 for scans using the wide (8 cm) collimation width at 100 and 120 kVp. For both tube voltages, VMI HU values tend to increase with increasing phantom size. The average accuracy over all insert materials was 1.3, 3.4, and 5.9 HU for the small, medium, and large phantom configuration at 100 kVp. The corresponding results at 120 kVp were 1.2, 2.2, and 4.5 HU. However, differences between both tube voltages were not statistically significant (*p* = 0.86). No difference between 4 and 8 cm collimation width was observed.

**Table 2.** Size dependence of the accuracy of virtual mono-energetic images (VMIs) for the small, medium, and large phantom configurations. The table lists mean error and standard deviation in Hounsfield units (HU). Results are reported for 100 and 120 kVp scans at a collimation width of 8 cm. Negative and positive errors indicate an under- and overestimation of the nominal value, respectively.

|  |  | Liver [HU] |  | Adipose [HU] |  | HA100 [HU] |  | HA400 [HU] |  |
| --- | --- | --- | --- | --- | --- | --- | --- | --- | --- |
|  |  | 100 kVp | 120 kVp | 100 kVp | 120 kVp | 100 kVp | 120 kVp | 100 kVp | 120 kVp |
| 40 keV | Small | 12.8±2.0 | 9.9±1.8 | 5.5±1.4 | 7.4±1.3 | 8.0±3.2 | 1.0±2.9 | -2.7±2.8 | -29.2±2.7 |
|  | Medium | 13.2±3.5 | 8.8±3.3 | 10.9±1.3 | 11.9±2.0 | 12.8±4.2 | 2.7±4.5 | -0.1±6.8 | -32.1±6.0 |
|  | Large | 20.2±5.1 | 14.0±4.7 | 15.5±4.6 | 16.1±3.1 | 19.3±8.8 | 7.4±6.7 | 10.0±16.1 | -28.4±14.4 |
| 50 keV | Small | 8.7±1.0 | 7.7±1.1 | 0.3±0.8 | 1.6±0.8 | 8.1±1.6 | 5.3±1.5 | 17.6±1.8 | 5.1±1.7 |
|  | Medium | 9.5±2.0 | 7.1±1.9 | 3.4±0.8 | 4.6±1.2 | 11.7±2.4 | 7.1±2.7 | 20.6±4.1 | 4.7±3.7 |
|  | Large | 13.8±2.9 | 10.9±2.7 | 6.2±2.7 | 6.9±1.8 | 15.6±4.9 | 10.1±3.9 | 27.5±9.4 | 8.0±8.6 |
| 60 keV | Small | 6.3±0.7 | 6.8±0.7 | -0.7±0.6 | 0.4±0.5 | 5.2±0.8 | 4.8±1.0 | 15.5±1.5 | 11.1±1.5 |
|  | Medium | 7.6±1.2 | 6.4±1.3 | 0.8±0.6 | 1.9±0.9 | 8.0±1.4 | 6.5±1.7 | 18.7±2.5 | 12.0±2.5 |
|  | Large | 10.3±1.6 | 9.4±1.6 | 2.6±1.7 | 3.4±1.2 | 10.4±2.7 | 8.5±2.4 | 23.6±5.6 | 15.2±5.3 |
| 70 keV | Small | 5.2±0.5 | 6.4±0.6 | -0.8±0.4 | 0.1±0.4 | 2.8±0.5 | 3.8±0.7 | 9.5±1.4 | 10.4±1.3 |
|  | Medium | 6.6±0.9 | 6.1±0.9 | -0.2±0.6 | 0.9±0.7 | 5.2±1.0 | 5.6±1.1 | 12.8±1.5 | 12.2±1.8 |
|  | Large | 8.3±1.2 | 8.6±1.1 | 1.0±1.2 | 1.8±1.0 | 6.8±1.7 | 7.2±1.6 | 16.4±3.2 | 15.1±3.3 |
| 100 keV | Small | 2.9±0.8 | 4.9±0.7 | -0.2±0.5 | 0.5±0.4 | -3.6±0.7 | -0.6±0.6 | -7.8±1.6 | -0.7±1.3 |
|  | Medium | 4.5±1.1 | 4.9±0.8 | -0.8±0.8 | 0.3±0.7 | -1.6±1.1 | 1.3±0.7 | -4.2±1.1 | 2.2±1.3 |
|  | Large | 5.0±1.3 | 6.8±0.8 | -0.5±1.1 | 0.5±0.9 | -1.3±1.6 | 2.1±1.2 | -2.0±1.6 | 5.1±1.3 |
| 150 keV | Small | 2.0±0.8 | 4.5±0.8 | 0.4±0.6 | 1.2±0.5 | -7.6±1.1 | -3.7±0.9 | -20.0±1.8 | -10.0±1.3 |
|  | Medium | 3.6±1.3 | 4.6±1.0 | -0.3±0.8 | 0.8±0.6 | -5.8±1.3 | -1.8±0.9 | -16.3±1.3 | -6.6±1.3 |
|  | Large | 3.3±1.6 | 5.9±1.0 | -0.5±1.3 | 0.4±1.0 | -5.9±2.2 | 1.3±1.5 | -14.8±2.3 | 3.9±1.5 |
| 200 keV | Small | 1.3±0.8 | 3.9±0.8 | 0.9±0.6 | 1.3±0.5 | -9.1±1.1 | -5.1±0.9 | -25.4±1.9 | -14.2±1.4 |
|  | Medium | 3.0±1.3 | 4.0±0.9 | -0.3±0.8 | 0.7±0.6 | -7.5±1.4 | -3.1±1.0 | -21.5±1.4 | -10.8±1.4 |
|  | Large | 2.9±1.8 | 5.7±1.0 | -0.6±1.3 | 0.3±1.0 | -7.8±2.5 | -2.6±1.7 | -20.2±2.5 | -8.2±1.6 |

A VMI of 62 keV was selected for further comparison, as it most closely matched the HU values obtained from the conventional 100 kVp data for all phantom sizes. Figure 2 shows the absolute HU difference from the average value over the three phantom sizes for conventional and 62 keV VMI data. For the medium phantom size (15 cm), both conventional and VMI provide a comparably low deviation of less than 3 HU. The VMI accuracy for the small and large phantom configuration is only slightly degraded. In contrast, conventional images demonstrate a substantially larger size-dependence with CT number variations of up to 28 HU.

**Figure 2.**
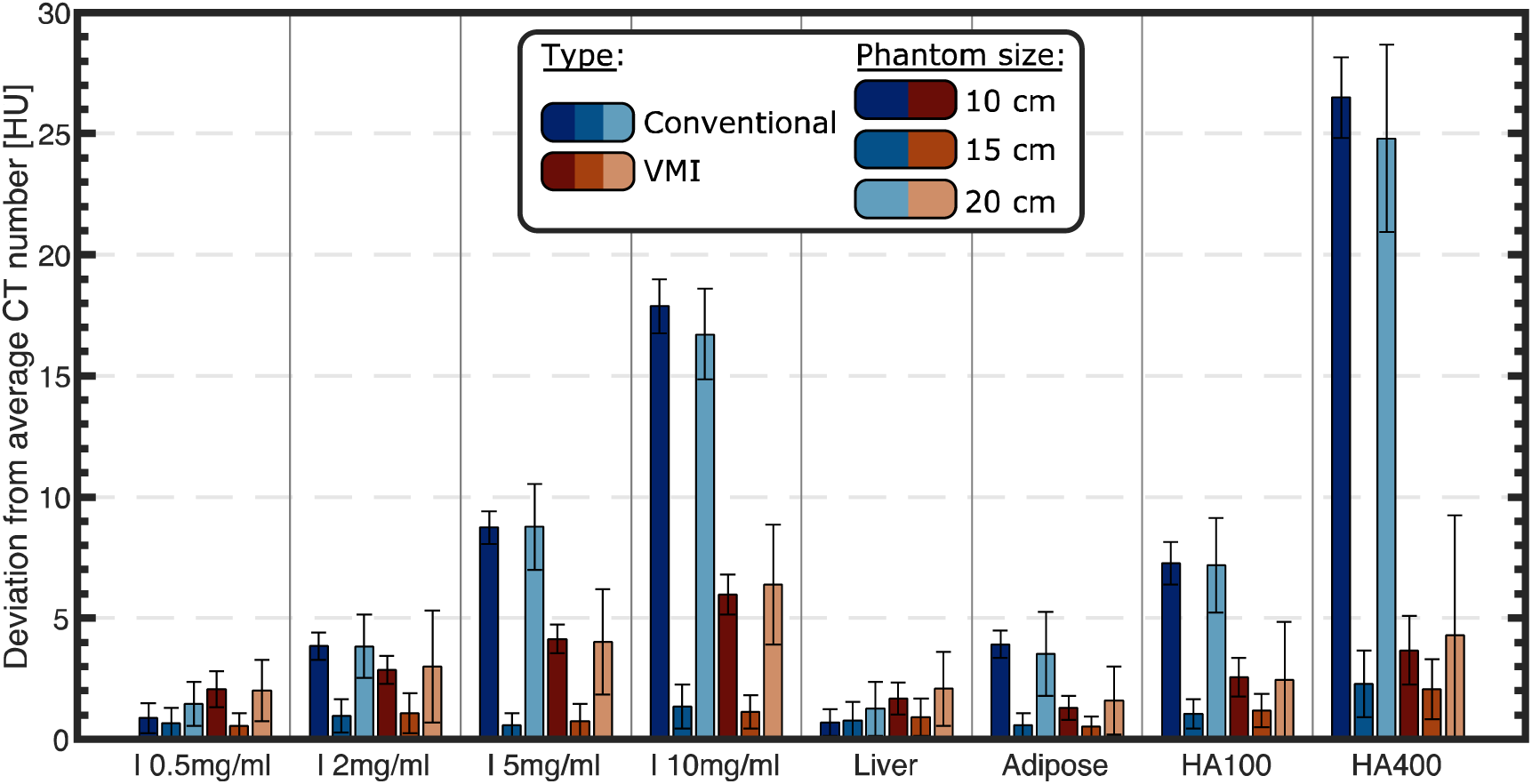
Bar plot of the (absolute) HU deviation from the size-averaged value for the conventional 100 kVp (blue) and the corresponding 62 keV virtual mono-energetic images (VMIs; red) data. The bars and whiskers represent the mean error and standard deviation, respectively. Different phantom sizes are displayed in different shades.

### 2. Iodine density

The error in quantified iodine density is shown in Figure 3 for inserts of increasing iodine concentration. A significant size dependence was observed for both tube voltages. Larger variations occurred for higher iodine concentrations and for smaller phantom geometries. 100 kVp yields slightly higher absolute iodine density errors of 0.17, 0.07, and 0.04 mg/ml compared to 120 kVp for the small, medium, and large phantom sizes, respectively. However, those differences were not significant for any configuration. Moreover, no significant difference in measurement error was found when comparing 4 and 8 cm collimation widths.

**Figure 3.**
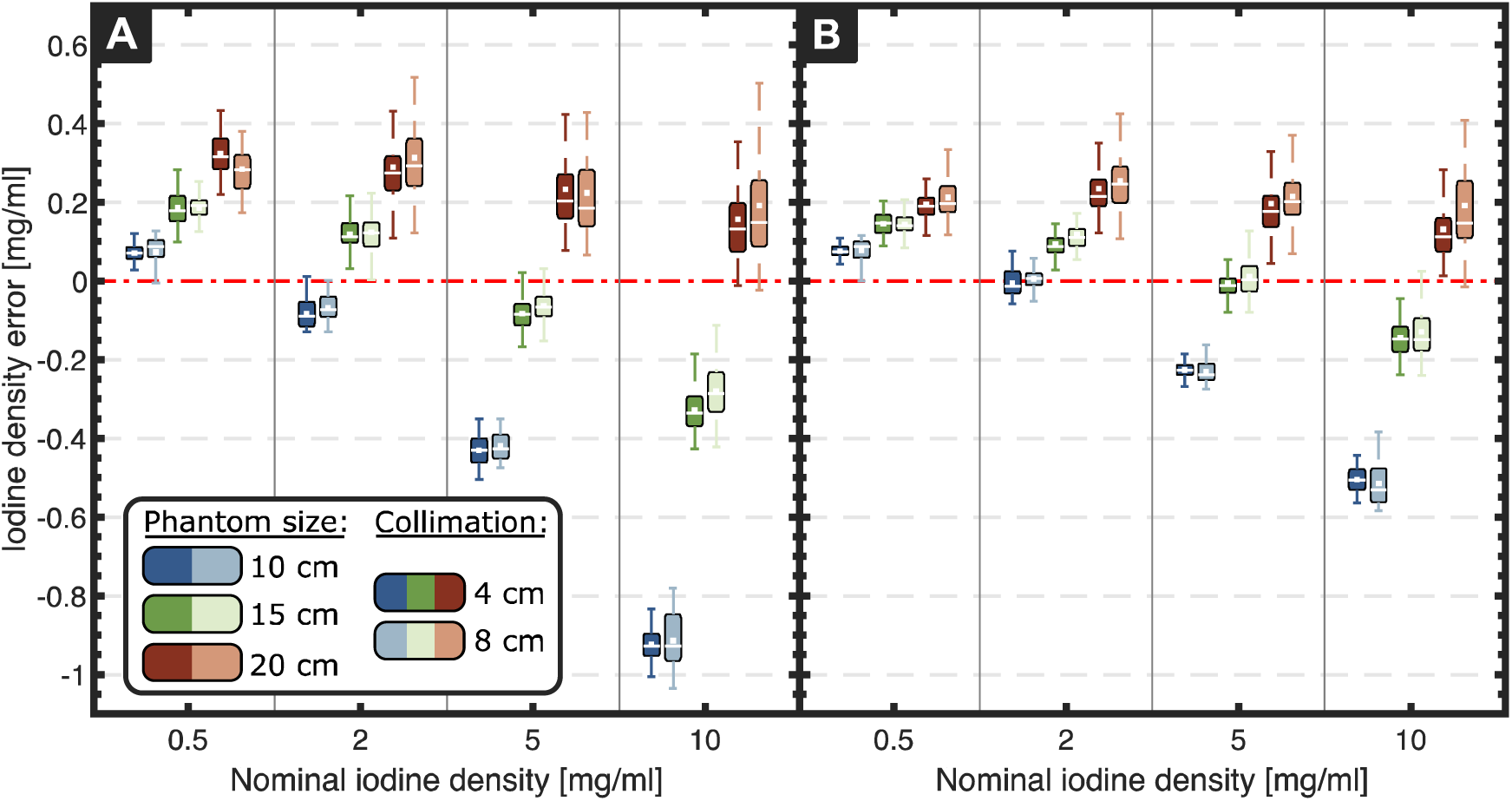
Iodine density errors for scans acquired at (A) 100 kVp and (B) 120 kVp. Mean and median of the distribution are represented by the central white line and marker, respectively, the box corresponds to the first and third quartile, and each whisker indicates 1.5 times the interquartile range. Different phantom sizes and collimation widths are displayed in different colors and shades, respectively.

### 3. Effective atomic number

Figure 4 shows the Z_eff_ errors obtained for the various acquisition and phantom configurations. Overall, the size-dependence is much smaller than for iodine density (Figure 3), except for adipose tissue. Both tube voltages showed no significant differences when comparing the measurement error among the three phantom sizes (100 kVp: *p* = 0.47, 120 kVp: *p* = 0.72). The mean accuracy was 0.23 and 0.16 for 100 kVp and 120 kVp scans, respectively. No significant difference between both collimation widths was observed.

**Figure 4.**
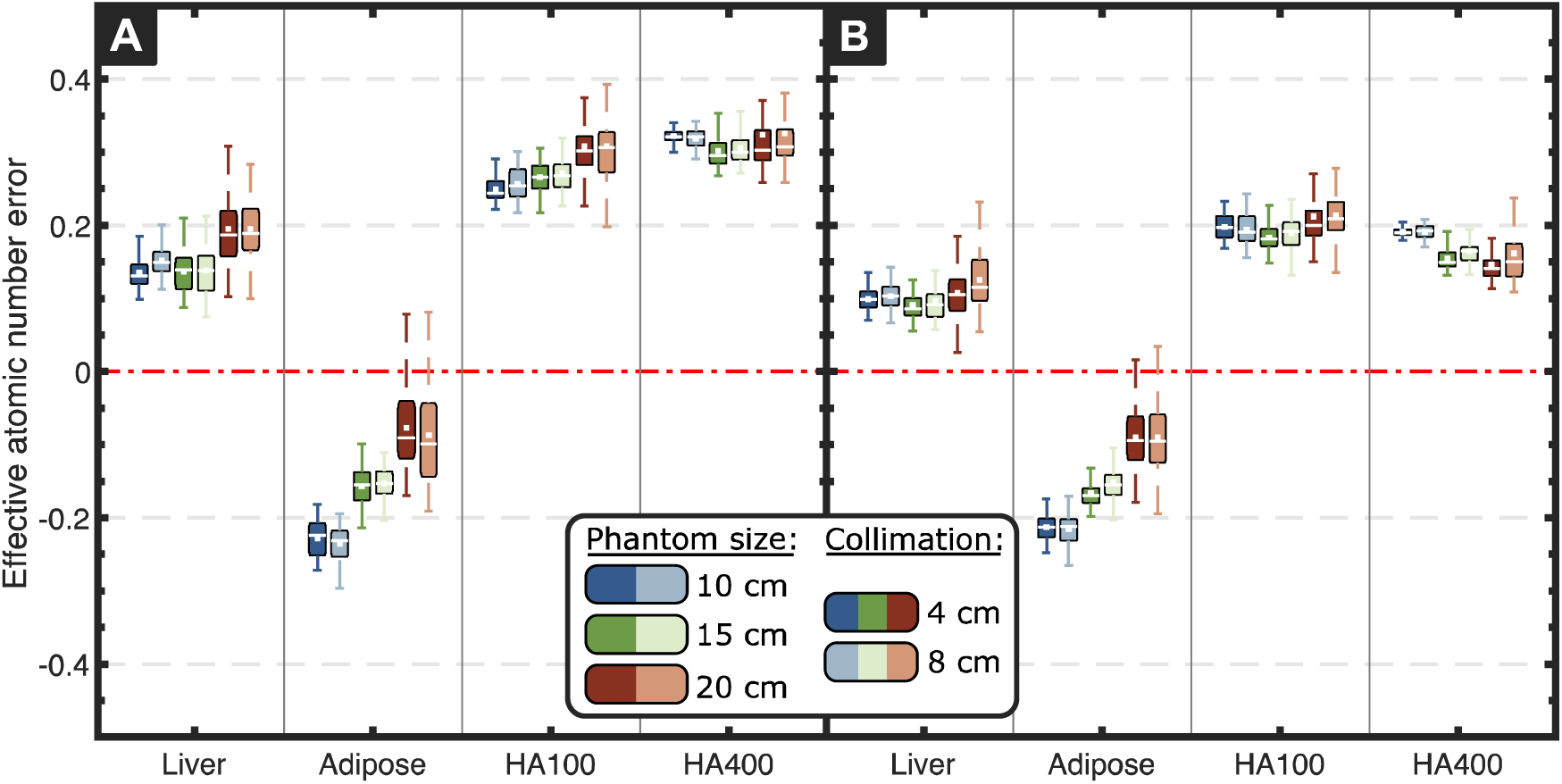
Effective atomic number (Z_eff_) errors for scans acquired at (A) 100 kVp and (B) 120 kVp. Mean and median of the distribution are represented by the central white line and marker, respectively, the box corresponds to the first and third quartile, and each whisker indicates 1.5 times the interquartile range. Different phantom sizes and collimation widths are displayed in different colors and shades, respectively.

### 4. Electron density

The electron density quantification is displayed in Figure 5. All mean errors were within the uncertainty of the nominal electron density values of 2 %ED_water_, except HA400 for the smallest phantom size at 100 kVp (2.1 %ED_water_ mean error). As for Z_eff_, there was no influence of phantom size on measurement error (100 kVp: *p* = 0.91, 120 kVp: *p* = 0.64). The mean electron density error was 0.72 and 0.34 %ED_water_ for 100 kVp and 120 kVp, with the largest differences occurring for HA400.

**Figure 5.**
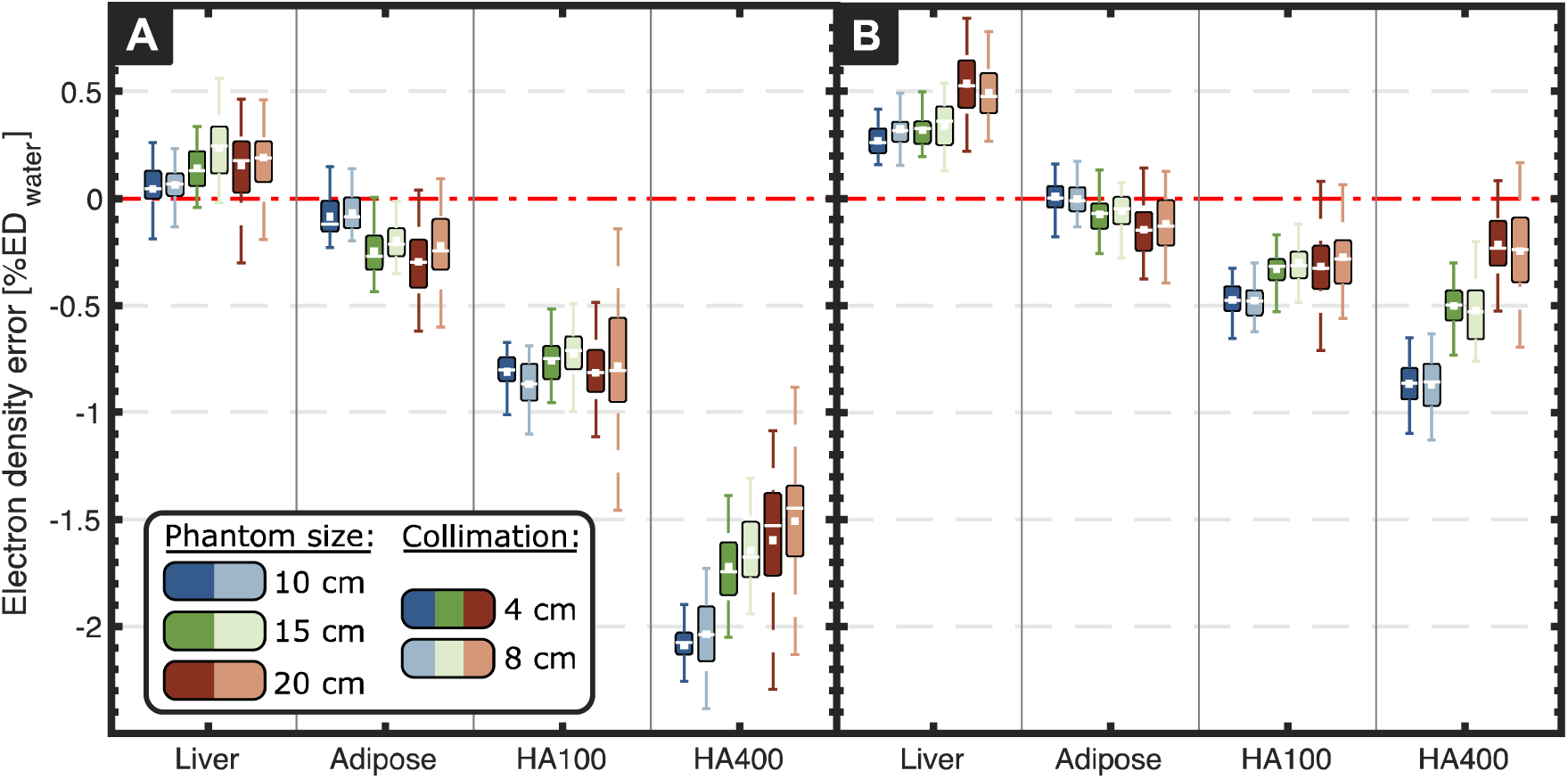
Electron density (ED) errors for scans acquired at (A) 100 kVp and (B) 120 kVp. The uncertainty of the nominal electron density values is 2 %ED_water_. Mean and median of the distribution are represented by the central white line and marker, respectively, the box corresponds to the first and third quartile, and each whisker indicates 1.5 times the interquartile range. Different phantom sizes and collimation widths are displayed in different colors and shades, respectively. No influence of the collimation on the electron density quantification was observed.

### 5. Radiation dose dependency

No significant differences in spectral accuracy (mean measurement errors) were found for both tube potentials and all phantom sizes when comparing results obtained at 0.9, 3, 6 and 9 mGy CTDI_vol_. Figure 6 shows the standard deviation of the spectral results obtained from all relevant insert materials. As expected, higher values are observed for larger phantom configurations and with lower imaging doses. However, the results obtained at 0.9 mGy were significantly different only for the 20 cm phantom. The corresponding standard deviations for 120 kVp scans were lower by 0.02 mg/ml, 0.01 and 0.05 %ED_water_, for iodine density, effective atomic number, and electron density, respectively. Significant differences between 4 (not shown) and 8 cm collimation widths were only observed for iodine density (100 kVp and 120 kVp) and electron density (120 kVp), and only with the smallest phantom configuration.

**Figure 6.**
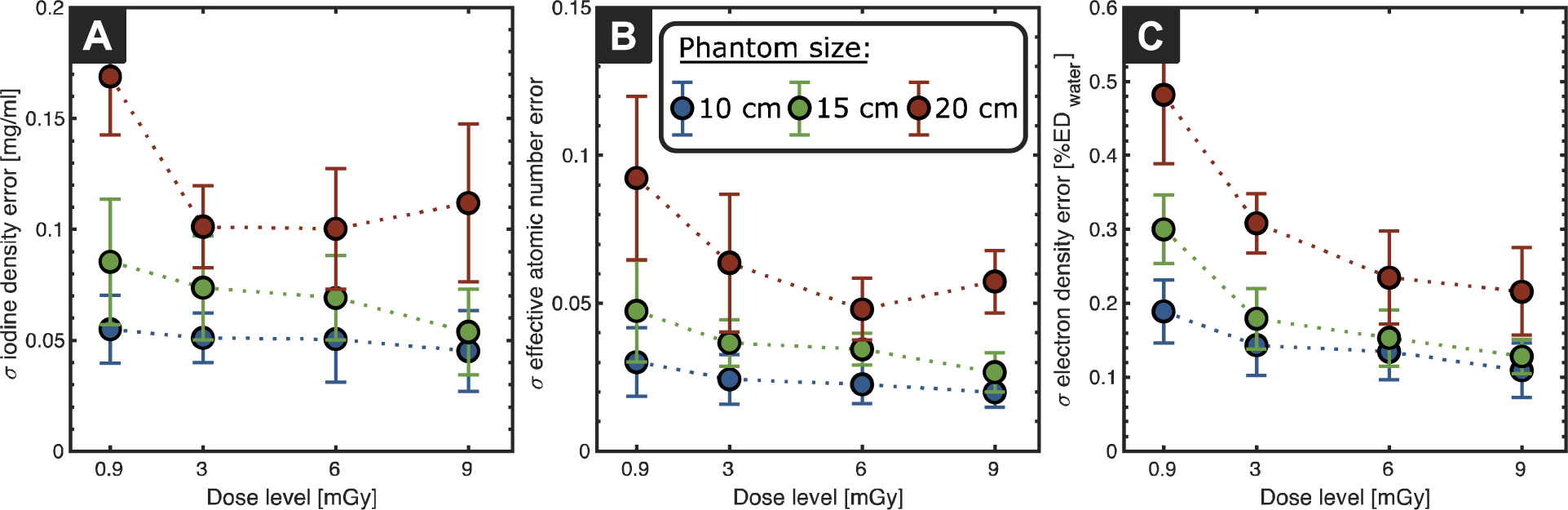
Impact of the dose level (CTDI_vol_; the nominal dose of the pediatric abdominal protocol is 9 mGy) on the standard deviation of spectral results: (A) iodine density, (B) effective atomic number, and (C) electron density obtained for scans at 8 cm collimation width.

## Discussion

While spectral imaging is becoming more widely utilized in clinical imaging, few studies have been published on DECT in the pediatric population and more experience is required for its adoption into clinical practice [14]. Precise and reliable spectral quantification is a key requirement of DECT. However, for pediatric imaging additional factors need to be considered including dose and scan time. While larger collimation width and lower tube voltage could be beneficial, they might compromise spectral quantification. We present the first investigation of quantitative spectral imaging performance at 100 kVp for a dedicated pediatric-sized phantom using the latest generation detector-based DECT.

62keV VMIs demonstrated very similar attenuation values (within few HUs) compared to conventional 100 kVp images across all insert materials for the medium phantom size (Figure 2). Moreover, VMIs substantially increased the stability of HU values for varying patient geometries, which can be attributed to accurate correction of beam hardening effects [23, 25]. Hence, VMIs can enable consistent attenuation quantification for pediatric patients despite the variability of body sizes occurring during childhood. This can improve the characterization and increase conspicuity of abdominal pathologies [16].

Iodine quantification was affected by the phantom size (Figure 3). Similar findings have been reported for previous generations of dual layer [23, 26] and dual source DECT [27]. Overall, measured iodine densities systematically increased with phantom size and changes were larger for higher iodine concentrations. However, the error was always within 1 mg/ml. Hence, radiologists need to consider patient size when interpreting images based on quantitative iodine density information. Future work should aim to improve the stability of material decomposition algorithms or develop adequate correction methods.

Effective atomic number and electron density results showed excellent accuracy (Figures 4 and 5) and were less sensitive to phantom size. This can be utilized to improve the relative stopping power accuracy in proton therapy treatment planning [28], which faces inherent limitations when relying on conventional CT images. Proton therapy allows sparing of normal tissues and reduces the integral dose, thereby making it very appealing for radiotherapy treatment in children [29].

Scans acquired at 120 kVp provided slightly better accuracy than 100 kVp, which can be attributed to the reduced spectral separation at lower tube potentials, as has been reported by others for 120/140 kVp [30, 31]. However, it should be noted that results depend on phantom size and attenuation [26]. The collimation width did not significantly affect spectral results (Figures 3-5). A larger collimation not only improves dose utilization but also reduces scan time, both crucial aspects for pediatric patients. It should also be noted that scans in this study were acquired with a very fast rotation time of 0.272 s.

Compared with its predecessor (IQon Spectral CT, Philips Healthcare), the overall spectral accuracy was improved [23]. For a comparable setting (medium phantom size, 9 mGy radiation dose, and 4 cm collimation width) the absolute error in VMI CT number, effective atomic number, and electron density are reduced by 8.69 HU, 0.03 and 0.45 %ED_water_, respectively, and the iodine density accuracy is comparable. The quantitative improvements between the two systems can be attributed to two technological advancements. First, the newer CT 7500 is equipped with a next-generation dual-layer x-ray detection system that is based on a higher-efficiency detector array coupled with a two-dimensional anti-scatter grid. In contrast, the previous generation detector array was based on a stick design that readout the scintillator from the side and necessitated a one-dimensional anti-scatter grid [10]. A second reason for the improvements in spectral quantification is an updated physics model of the detector system that is utilized in the 2-base material decomposition process. These improvements include more accurate detector design inputs and the inclusion of second-order physics effects, i.e., light-matter interactions, that occur within the detector system.

To the best of our knowledge there are no dedicated studies comparing the imaging performance of different DECT implementations for pediatric imaging. While many have compared the imaging performance of different DECT technologies in larger phantoms [32, 33], results might differ for pediatric-sized phantoms. Moreover, the recent clinical implementation of the first commercial photo-counting CT [34–36] may open new possibilities for spectral imaging that can benefit pediatric patients; however, future studies are needed to investigate and compare the pediatric imaging performance.

The findings of our work evidence the high image quality achievable using standard pediatric abdominal protocol doses. The use of lower tube voltage and dedicated CT protocols are critical to avoid high effective doses due to the smaller transverse body diameter in children, which highlights the need for a careful trade-off between dose and image quality [37]. In fact, it has been demonstrated that DECT scans of the head and abdomen in children can be dose-neutral compared to single energy CT [38, 39]. We found that spectral quantification was robust with further dose reductions down to 10% of the protocol’s nominal value (Figure 6). Variations were slightly higher for larger phantom sizes due to their increased attenuation, which is in line with previous publications [23, 31].

A limitation of our study is the use of a geometric phantom. This enables consistent evaluation of spectral results with known ground truth but does not include anatomical features or organs. Therefore, future investigations should be extended to more realistic scenarios such as pediatric anthropomorphic or 3D printed [40] phantoms (ideally tailored to DECT applications). We also only employed a single scan protocol. However, results are not expected to vary much with scan parameters due to the simultaneous acquisition of both spectra [23].

Spectral imaging enables characterization of materials based on their attenuation at different energies and has resulted in various novel applications of CT imaging within the last two decades. Our initial phantom evaluation has shown that detector-based DECT imaging at 100 kVp maintains a high quality of spectral accuracy. Even though DECT offers several clear advantages, further clinical studies are needed for the broad integration of spectral information into pediatric imaging due to its unique requirements.

## Data Availability

All data produced in the present study are available upon reasonable request to the authors.

## Acknowledgment

The authors acknowledge support through the National Institutes of Health (R01EB030494). In addition, the authors would like to thank Michael Geagan for his help with 3D-printing the extension rings and Safaa Abdallah for her help with acquiring the data.

## Conflict of interest

Harold I. Litt and Peter B. Noël received a hardware grant from Philips Healthcare. Peter B. Noël receives research grant funding from Philips Healthcare. Dr. Halliburton is an employee of Philips Healthcare. The other authors have no relevant conflicts of interest to disclose.

## Notes

### Competing Interest Statement

Harold I. Litt and Peter B. Noel received a hardware grant from Philips Healthcare. Peter B. Noel receives research grant funding from Philips Healthcare. Dr. Halliburton is an employee of Philips Healthcare. The other authors have no relevant conflicts of interest to disclose.

